# Interleukin-6 receptor genetic variation and tocilizumab treatment response to COVID-19

**DOI:** 10.1101/2021.04.24.21256047

**Authors:** Ammar Ali Almarzooq

## Abstract

Interleukin-6 receptor (IL6R) stimulates the inflammatory pathways as part of the acute-phase response to infection. Tocilizumab is a monoclonal antibody that inhibits both membrane-bound and soluble IL6R and is used to treat inflammatory conditions, including COVID-19. Despite the disproportionate incidence of COVID-19 among underserved, racial, and ethnic minority populations, the efficacy of tocilizumab in hospitalized COVID-19 patients from these populations is unclear. In this work, three genetic markers for the *IL6R* gene were analyzed across diverse ethnic backgrounds to identify population differences in response to tocilizumab treatment. Genetic structure analyses showed that African populations were significantly different from other described populations. In addition, mapped frequencies of these alleles showed that Sub-Saharan African populations were 3.4x more likely to show an impaired response to tocilizumab than East Asian populations, and 1.8x more likely than European ancestry populations. Existing *IL6R* genotype results may identify populations at increased therapeutic failure risk. As results from current clinical trials on the efficacy of tocilizumab treatment for extreme COVID-19 infections are conflicting, more studies are needed across diverse patient backgrounds to better understand the genetic factors necessary to predict treatment efficacy. This work demonstrates how pharmacogenomics studies can elucidate genetic variation on treatment efficacy on COVID-19.

## INTROUCTION

Over 150 million cases and 3.1 million deaths have occurred world-wide as a result of the coronavirus disease 2019 (COVID-19) pandemic [1]. Treatment options are still limited, with only glucocorticoids known to improve survival rates among severely ill patients [2] by alleviating the excessive host inflammatory response [3].

Interleukin-6 receptor (IL6R) is released in response to infection and stimulates the inflammatory pathways as part of the acute-phase response. Tocilizumab, a monoclonal antibody that inhibits membrane-bound and soluble interleukin-6 receptors, is used to treat inflammatory conditions such as rheumatoid arthritis and cytokine release syndrome following cell therapy. Clinical use has been described in COVID-19 [4-6], and following successful trials, tocilizumab has now become a standard National Health Service NHS treatment for specific COVID-19 patients [7]. In addition, The Infectious Diseases Society of America (IDSA) has backed the use of tocilizumab for COVID-19 treatment [8]. However, results from clinical trials using this treatment are mixed, with specific trials finding no benefit of the treatment among patients with severe disease [9-12]. In one case, tocilizumab treatment made no significant difference in clinical status or mortality rate even after 28 days among hospitalized patients with severe COVID-19 pneumonia [10]. However, another independent study demonstrated that survival improved ∼70% for patients treated with tocilizumab (or sarilumab), relative to standard care alone [3]. This is consistent with another U.K. based study showing a slight, but significant, mortality benefit for critically ill COVID-19 patients treated with tocilizumab [13]. Comparisons across these studies is misleading, however, due to varying sample sizes, demographics, and methodology.

Genetic factors are one of the major contributors to individual and ethnic differences in drug therapeutic efficacy and toxicity [14-15]. Therefore, medication dosing might need to be altered based on a patient’s genetic information [16-17]. There are several gene variants that can alter metabolism and processing of tocilizumab, potentially increasing the risk of therapeutic failure [18-19]. Patients with *IL6R* rs4329505 CC and CT genotypes, may have decreased response to tocilizumab when compared to patients with TT genotype [18]. Also, rs12083537 AA genotype is associated with decreased response to tocilizumab and higher risk for asthma compared to AG genotype [18]. Finally, rs11265618 CC genotype is associated with an increased response to tocilizumab when compared to CT and TT genotypes [19].

The goal of this study was to use pharmacogenetic and modeling based approaches to better understand, and potentially predict, genetic factors related to treatment efficacy of tocilizumab. Such results may elucidate the underlying genetic factors and resolve the conflicting reports from COVID-19 clinical trials treating with tocilizumab. We show that pharmacogenomics can enhance our understanding of COVID-19 treatment failure and support advancement of the drug development pipelines.

## RESULTS

### Genetic structure of IL6R across populations

Three variants across 26 global populations from the 1000 Genomes Project Phase III (1kG-p3) dataset were selected (TAB. S1). SNPs were tested for Hardy–Weinberg equilibrium (HWE), and populations included in this study met HWE standards (p > 0.05) across the three SNPs, except for the Esan in Nigeria population (ESN) (p = 0.039) (TAB. S1). rs4329505 was in pair-wise linkage disequilibrium (LD) with rs11265618 (r2 ≥ 0.90) in European (EUR) and East Asian (EAS) populations.

The two leading principal components from the three variants, showed well-defined separation between African (AFR) populations and other global populations (FIG. 1a). This was further validated using pairwise F_ST_ analyses (TAB. S2). The lowest level of differentiation was observed between EUR and South Asian (SAS) populations, followed by East Asian and American (AME) populations. The greatest affinity with AFR was observed with EUR and South Asian populations (FIG. 1b).

**Figure 1:**
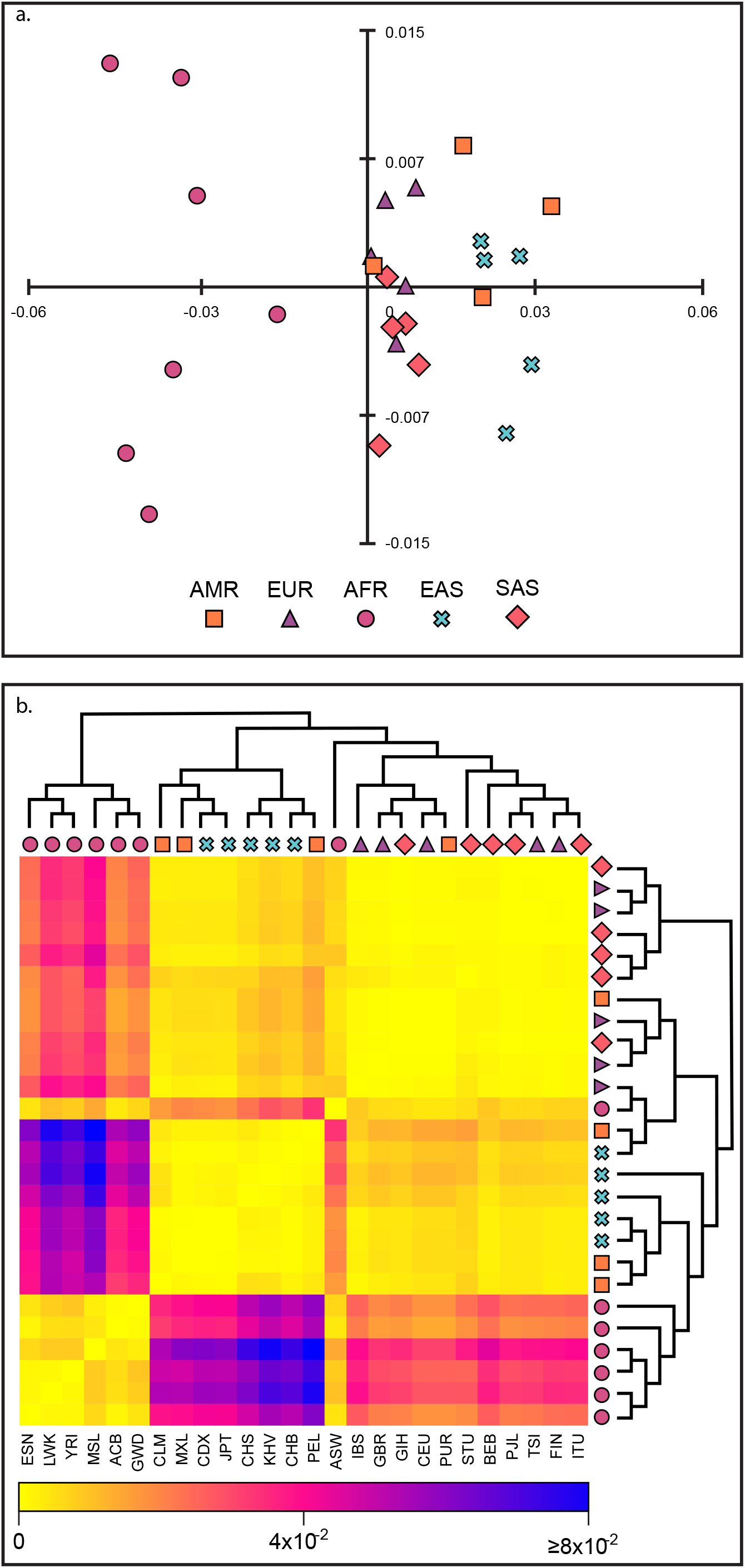
Population structure analysis. **a**. MDS plot of the three variants across 26 global populations from the 1000 Genomes Project Phase III (1kG-p3) dataset. **b**. Heat map showing pairwise F_ST_ values with hierarchical clustering. Yellow for the lowest and blue for the highest F_ST_ values. All the abbreviations names and detail information of the reference populations are in TAB. S1 and S2.

### Pharmacogenetic analyses by biogeographic grouping system

Across the nine biogeographical groups, 80% of subjects were of EUR, Sub-Saharan African (SSA), South Central Asian, and East Asian origin (20% each), with Latino (LAT) and African Americans/Afro-Caribbeans (AAC) comprising the remaining 20% (14% and 6%, respectively) (TAB. 1). SNPs were tested for Hardy–Weinberg equilibrium (HWE), and all the studied biogeographical groups fulfilled HWE (p > 0.05) (TAB. 1).

Distinct differences were found among these populations, with direct impacts to tocilizumab clinical outcomes (FIG. 2). rs12083537 allele frequency was significantly higher for SSA (0.337), followed by AAC (0.295), EUR (0.193), LAT (0.166) and South Central Asians (0.158) indicating a decreased metabolism and clearance of tocilizumab when compared to East Asians (0.110) (TAB. 1). rs11265618 allele frequency was significantly higher in SSA (0.347), followed by AAC (0.291), South Central Asians (0.194), EUR (0.186) and LAT (0.128) indicating a decreased metabolism and clearance of tocilizumab when compared to East Asians (0.104) (TAB. 1). Suggesting that SSA populations are 3.4x more likely to show impaired response to tocilizumab than East Asian populations, and 1.8x more likely than EUR populations (FIG. 3 and TAB. 1). The rs4329505 marker was excluded from the analysis due to contradictory genotype-phenotype studies toward tocilizumab (FIG. 2) (18,19).

**Figure 2:**
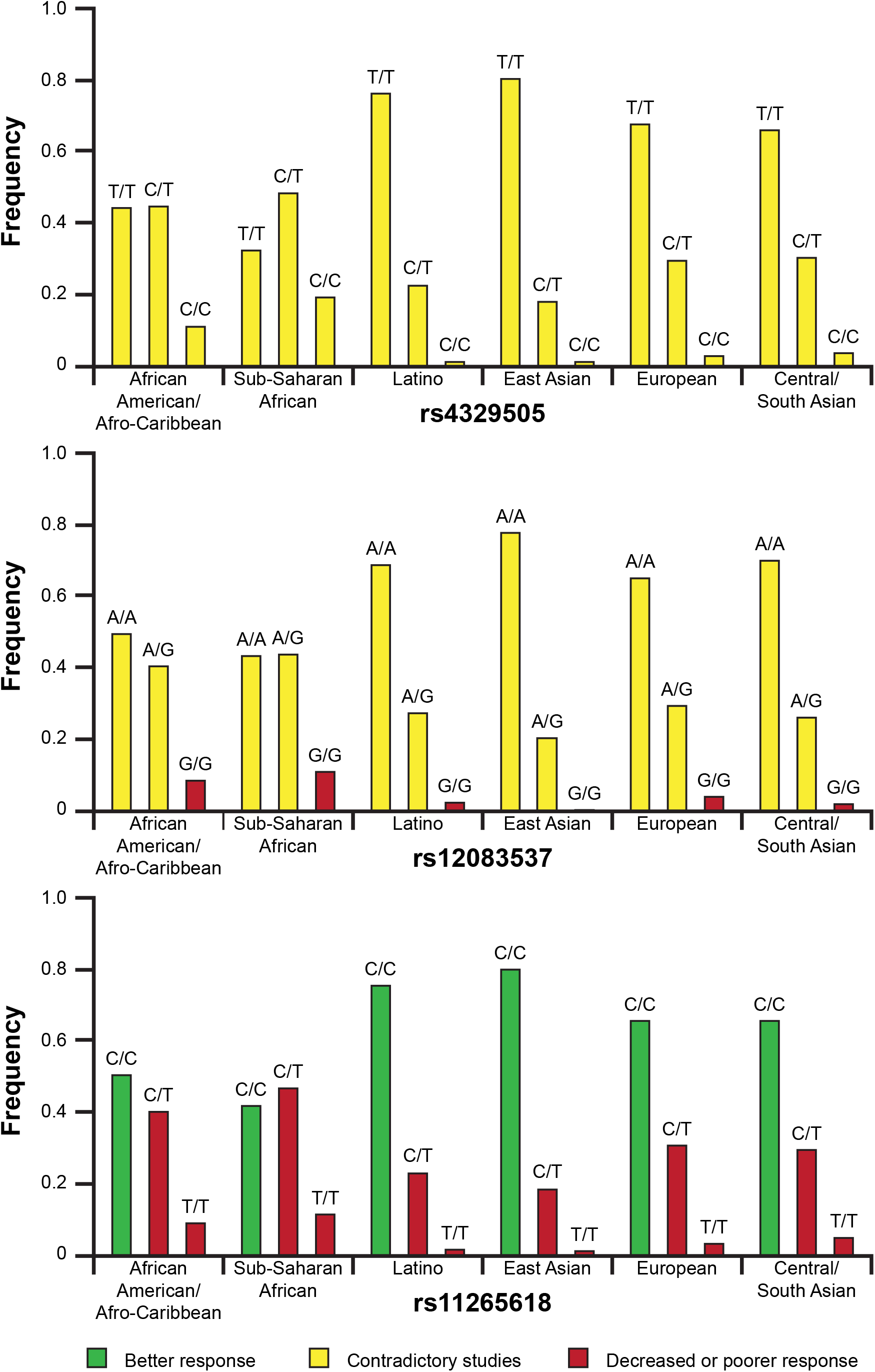
Biogeographical groups and pharmacogenetic analyses for *IL6R* and tocilizumab associations. Frequencies of three pharmacogenetics (PGx) genotypes and phenotypes assessed cumulatively against the nine biogeographical groups.

**Table 1:**
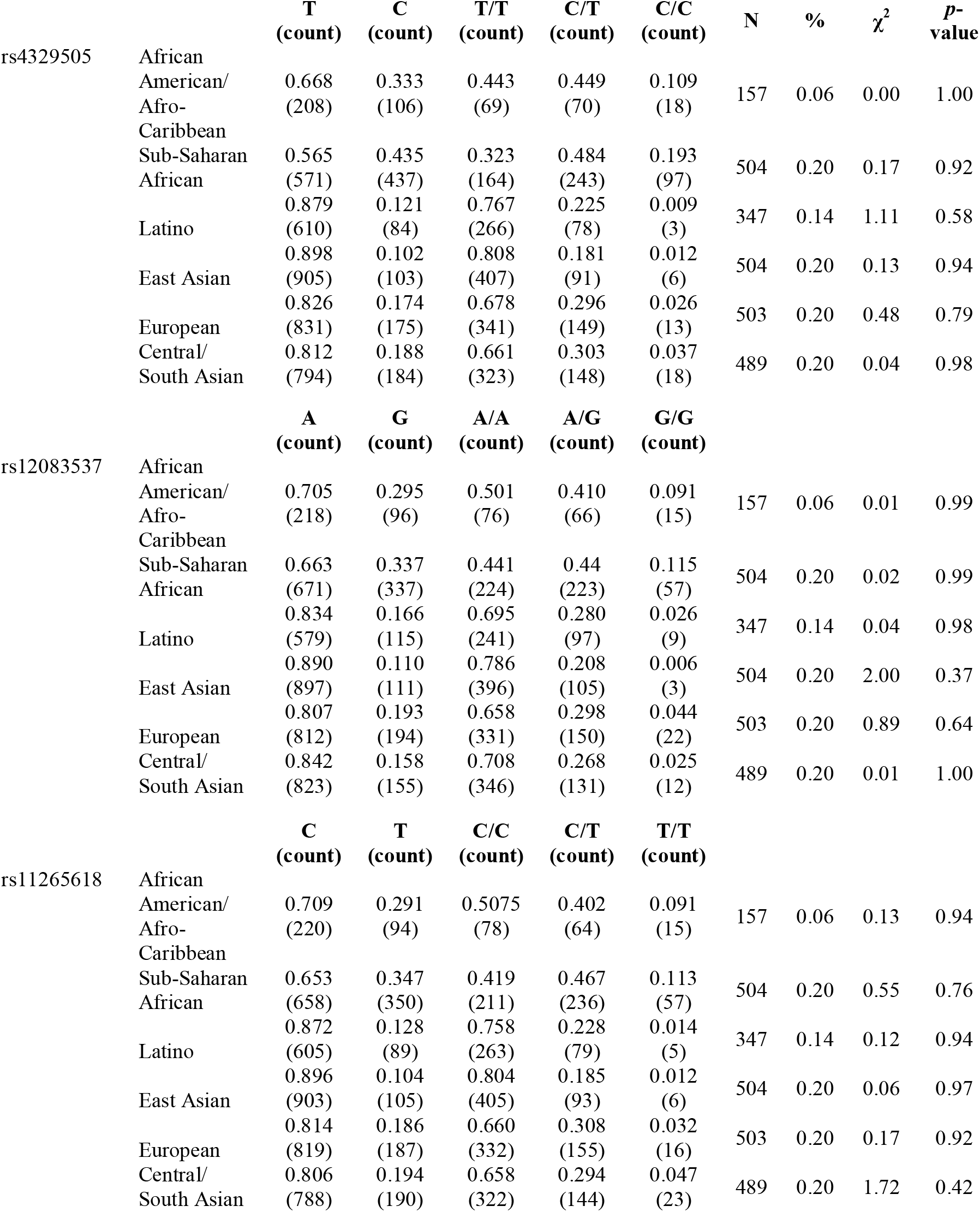
Distribution of *IL6R* alleles and genotypes assessed cumulatively against the biogeographical groups (rs4329505, rs12083537, rs11265618)

**Figure 3:**
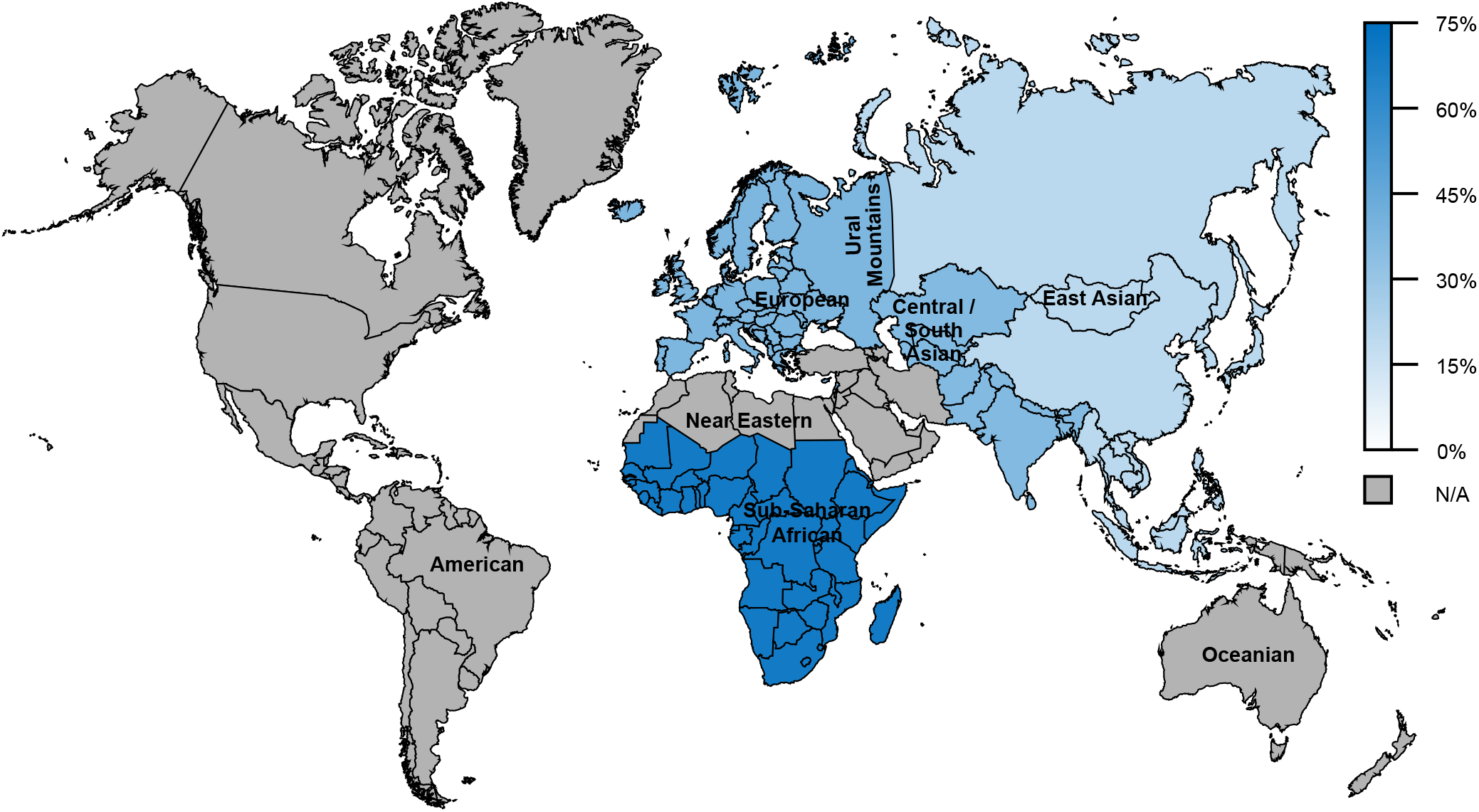
Global frequency map of variability in of *IL6R* reduced response phenotypes on tocilizumab response. Since this map indicates the borders of each geographical group based on the location of genetic ancestors pre-Diaspora and pre-colonization, the two admixed groups (AAC and LAT) are absent. It should also be noted that, due to the large geographical areas designated for each group, a single population may not accurately capture the large amount of genetic diversity found in that defined region.

## DISCUSSION

Despite the disproportionate incidence of COVID-19 infection among racial and ethnic minority populations [20-28], data regarding the safety and efficacy of repurposed treatments for patients within these populations are lacking [29]. In addition, conflicting results of the use of these treatment in clinical trials warrants further investigation. *IL6R* genotyping may help identify populations at an increased risk of experiencing ADRs or therapeutic failure as a result of tocilizumab treatment. However, the role of race and ethnicity on the clinical course of COVID-19 is complex due to the potential interplay of genetics, social determinants, socioeconomic factors, and historical and structural inequities[23, 25, 28, 30].

In this work, several genetic markers were analyzed across diverse ethnic backgrounds to identify population level differences in drug responses may be associated with tocilizumab treatment. MDS analysis showed that the African populations significantly deviated from other populations. MDS results were further validated by pairwise F_ST_ values (FIG. 1). Mapped frequencies of these alleles showed that Sub-Saharan African populations were 3.4x more likely to show impaired *IL6R* than East Asian populations, and 1.8x more likely than European ancestry populations (FIG. 3 and TAB. 1). Therefore, it is plausible that a higher proportion of Asian and European ancestry populations could have a “normal” tocilizumab response, and therefore may be less susceptible to complications from tocilizumab-based treatment. Our analysis excluded rs4329505 marker not only due to conflicting research on genotype-phenotype for tocilizumab treatment (FIG. 2) [19, 20], but also because the pair-wise LD with rs11265618 (r2 ≥ 0.90) in Europeans and East Asian populations. This is consistent with the recently updated PharmGKB Clinical Annotations change downgrading Level of Evidence from three to four [31]. However, polymorphisms in other genes may also be associated with tocilizumab response. Patients with the *CD69 rs11052877* AA genotype, for example, can have an increased response to tocilizumab compared to patients with the AG and GG genotypes [32]. Also, patients with the *FCGR3A rs396991 AA* genotype may have an increased response to tocilizumab as compared with patients with the AC or CC genotypes [33]. Finally, *GALNT18 rs4910008* CC individuals may also have an increased response to tocilizumab compared with patients with the CT and TT genotypes [32].

Results from this study show that pharmacogenomics studies can enhance our understanding of adverse reactions to COVID-19 treatments and support advancement of drug development pipelines.

## METHODS

### Population structure analyses

Population structure was examined using Wright’s Fixation Index (F_ST_) for pairwise distances between populations based on three SNPs across *IL6R* to identify the ancestral relatedness across 26 global populations from the 1000 Genomes Project Phase III (1kG-p3) dataset [34]. Ethnically defined populations by the 1kG-p3 dataset consisted of 2504 individuals from 26 populations within five defined ancestral groups (TAB. S1):

1. African (AFR): Americans of African Ancestry in SW, USA (ASW), Esan in Nigeria (ESN), Gambian in Western Divisions in the Gambia (GWD), Mende in Sierra Leone (MSL), Luhya in Webuye, Kenya (LWK), Yoruba in Ibadan, Nigeria (YRI) and African Caribbeans in Barbados (ACB).
2. European (EUR): Utah Residents (CEPH) with Northern and Western European Ancestry (CEU), Finnish in Finland (FIN), British in England and Scotland (GBR), Iberian Population in Spain (IBS) and Toscani in Italia (TSI).
3. East Asian (EAS): Han Chinese in Beijing, China (CHB), Chinese Dai in Xishuangbanna, China (CDX), Southern Han Chinese (CHS), Japanese in Tokyo, Japan (JPT) and Kinh in Ho Chi Minh City, Vietnam (KHV).
4. South Asian (SAS): Bengali from Bangladesh (BEB), Indian Telugu from the UK (ITU), Punjabi from Lahore, Pakistan (PJL), Tamil from the UK (STU) and Gujarati Indian from Houston, Texas (GIH).
5. American (AMR): Mexican Ancestry from Los Angeles USA (MXL), Puerto Ricans from Puerto Rico (PUR), Colombians from Medellin, Colombia (CLM) and Peruvians from Lima, Peru (PEL).

Genotype and allele frequencies were tested for deviations from Hardy-Weinberg equilibrium using a chi-square (χ^2^) test (p □ > □ 0.05). These were implemented with the HWChisq function in HardyWeinberg R package [35]. Fixation indices were estimated with the calcFst function in the polysat R package [36]. Divergences were visualized by multidimensional scaling analysis (MDS) using the metaMDS function in the vegan R package [37] and with a hierarchical clustering based on F_ST_ values using the hclust function in base R.

### Pharmacogenetic analyses

Frequencies of the three pharmacogenetics biomarkers were assessed cumulatively for nine biogeographical groups [38], that consisted of 2,504 individuals from 26 global populations [34].

These nine groups were defined by global autosomal genetic structure and based on data from large-scale sequencing initiatives, and are used to illustrate the broad diversity of global allele frequencies in this study. Genotype and allele frequencies were tested for deviations from Hardy-Weinberg equilibrium using a χ^2^ test (p□>□0.05). These were implemented with the HWChisq function in HardyWeinberg R package [35. LD analysis was performed using the LDlink tool to generate r^2^ values [39]. This biogeographic grouping system meets a key need in pharmacogenetics research by enabling consistent communication of the scale of variability in global allele frequencies and is now used by PharmGKB and CPIC [38].

1. American (AME): The American genetic ancestry group includes populations from both North and South America with ancestors predating European colonization, including American Indian, Alaska Native, First Nations, Inuit, and Métis in Canada, and Indigenous peoples of Central and South America.
2. Central/South Asian (SAS): The Central and South Asian genetic ancestry group includes populations from Pakistan, Sri Lanka, Bangladesh, India, and ranges from Afghanistan to the western border of China.
3. East Asian (EAS): The East Asian genetic ancestry group includes populations from Japan, Korea, and China, and stretches from mainland Southeast Asia through the islands of Southeast Asia. In addition, it includes portions of central Asia and Russia east of the Ural Mountains.
4. European (EUR): The European genetic ancestry group includes populations of primarily European descent, including European Americans. We define the European region as extending west from the Ural Mountains and south to the Turkish and Bulgarian border.
5. Near Eastern (NEA): The Near Eastern genetic ancestry group encompasses populations from northern Africa, the Middle East, and the Caucasus. It includes Turkey and African nations north of the Saharan Desert.
6. Oceanian (OCE): The Oceanian genetic ancestry group includes pre-colonial populations of the Pacific Islands, including Hawaii, Australia, and Papua New Guinea.
7. Sub-Saharan African (SSA): The Sub-Saharan African genetic ancestry group includes individuals from all regions in Sub-Saharan Africa, including Madagascar.
8. African American/Afro-Caribbean (AAC): Individuals in the African American/Afro-Caribbean genetic ancestry group reflect the extensive admixture between African, European, and Indigenous ancestries and, as such, display a unique genetic profile compared to individuals from each of those lineages alone. Examples within this cluster include the Coriell Institute’s African Caribbean in Barbados (ACB) population, the African Americans from the Southwest US (ASW) population, and individuals from Jamaica and the US Virgin Islands.
9. Latino (LAT): The Latino genetic ancestry group is not defined by an exclusive geographic region, but includes individuals of Mestizo descent, individuals from Latin America, and self-identified Latino individuals in the United States. Like the African American/Afro-Caribbean group, the admixture in this population creates a unique genetic pattern compared to any of the discrete geographic regions, with individuals reflecting mixed native and indigenous American, European, and African ancestry.

The total frequency of the three SNPs within these nine geographically-defined groups were mapped for global impact visualization of allele frequency on tocilizumab response (TAB. 1, FIG. 2). Inferred frequencies for contradictory genotype-phenotype were excluded from our biogeographical analyses.

## Data Availability

The datasets generated during the current study are included in the supplementary files.

## ACKNOWLEDGEMENTS

N/A

## AUTHORS’ CONTRIBUTIONS

AA conceived the research study, performed all the analyses reported and wrote the manuscript.

## CONSENT FOR PUBLICATION

N/A

## COMPETING INTERESTS

The author declares that there is no conflict of interest.

## DATA AVAILABILITY

The datasets generated during the current study are included in the supplementary files.

## ETHICS DECLARATIONS

N/A

## Notes

### Competing Interest Statement

The authors have declared no competing interest.

